# Hospitals That Report Severe Sepsis and Septic Shock Bundle (SEP-1) Compliance Have More Structured Sepsis Performance Improvement

**DOI:** 10.1101/2021.05.11.21257054

**Authors:** Ty B. Bolte, Morgan B. Swanson, Anna Kaldjian, Nicholas M. Mohr, Jennifer McDanel, Azeemuddin Ahmed

## Abstract

**Objective:** Sepsis is a common cause of death in hospitalized patients. The Centers for Medicare & Medicaid Service (CMS) Severe Sepsis and Septic Shock Bundle (SEP-1) is an evidence-based early management bundle focused on improving sepsis outcomes. It is unknown which quality improvement (QI) practices are associated with SEP-1 compliance and if those practices reduce sepsis mortality. The objectives of this study were to compare sepsis QI practices in SEP-1 reporting and non-reporting hospitals and to measure the association between specific elements of sepsis QI processes and SEP-1 performance and hospital-specific risk-adjusted sepsis mortality.

**Design, Setting, and Patients:** This mixed methods study linked telephone survey data on QI practices from Iowa hospitals to SEP-1 performance data and risk-adjusted mortality from statewide all-payer administrative claims database. The survey assessed sepsis QI practices in eight categories. Characteristics of hospitals and sepsis QI practices were compared by SEP-1 reporting status. Univariable and multivariable logistic and linear regression estimated the association of QI practices with hospital SEP-1 performance and observed-to-expected sepsis mortality ratios.

**Interventions:** None

**Measurements and Main Results:** All 118 Iowa hospitals completed the survey (100% response rate). SEP-1 reporting hospitals were more likely to have sepsis QI practices, including reporting sepsis quality to providers (64% vs. 38%, p = 0.026) and using the case review process to develop sepsis care plans (87% vs. 64%, p = 0.013). Sepsis QI practices were not associated with increased SEP-1 scores. Two were associated with reduced mortality: having a sepsis committee β= -0.11, p = 0.036) and using case review results for sepsis care plans (β= -0.10, p = 0.049).

**Conclusions:** Hospitals reporting SEP-1 compliance to CMS conduct more sepsis QI practices. Most QI practices are not associated with increased SEP-1 performance or decreased sepsis mortality. Future work could explore how to implement these performance improvement practices in hospitals not reporting SEP-1 compliance.

## Introduction

Each year, approximately 1.7 million adults are diagnosed with sepsis, resulting in more than 250,000 deaths. Health care costs in the U.S. due to sepsis are estimated to be over $62 billion annually. (1), (2) Performance improvement activities have been a hallmark of sepsis care since the Surviving Sepsis Campaign developed sepsis bundles and multidisciplinary quality improvement programs many years ago. In addition, performance improvement programs aimed at improving sepsis bundle treatment have demonstrated marked reduction in sepsis mortality even after adjustment for comorbidities, age, and disease severity. (3) Other studies have confirmed the benefits of using “sepsis bundles” to improve patient outcomes.(4-6) Based on this work, the Centers for Medicare & Medicaid Services (CMS) adopted the Early Management Bundle, Severe Sepsis/Septic Shock (SEP-1) quality measure in 2015.(6, 7) Since its introduction, the SEP-1 measure has drawn attention to sepsis quality and focused the efforts of hospital quality departments on sepsis performance. (8)

CMS measures provide process metrics that are commonly used for quality measurement and improvement in a variety of conditions, including heart failure, osteoporosis, stroke, and myocardial infarction. (9-13) Many hospitals have used these initiatives to narrow the focus of disease-specific quality initiatives, but a recent survey of hospital quality officers found that the complexity of SEP-1 was an obstacle in meeting the requirements of this measure.(14) This complexity makes SEP-1 an important focus not only for improving sepsis outcomes, but also for understanding how quality measurement impacts the structure and process of performance improvement.

The objectives of this study are to (1) compare sepsis performance improvement activities in hospitals that do and do not report SEP-1 performance and (2) to identify elements of sepsis performance improvement associated with better SEP-1 scores and outcomes. Our goal is to identify elements of sepsis performance improvement processes associated with better SEP-1 scores and outcomes to inform hospital performance improvement programs.

## Methods

### Study Setting and Design

This study was a mixed methods analysis linking hospital performance improvement activities measured from a telephone survey of hospital sepsis coordinators with CMS SEP-1 reporting data and clinical outcomes measured using all-payer administrative claims in a rural, Midwestern state. Our survey of quality improvement and safety coordinators at all acute care hospitals in the state was completed between May 2020 to July 2020. Outcome data for sepsis bundle adherence and hospital aggregate sepsis mortality were linked from publicly available hospital quality reporting data and risk-adjusted mortality calculated from statewide all-payer inpatient and ED administrative claims data. This study was determined not to be human subjects research by the local Institutional Review Board and is reported according to the STrengthening the Reporting of OBservational studies in Epidemiology (STROBE) guidelines.(15)

### Survey of Hospital Performance Improvement Practices

A telephone survey of hospital quality improvement and safety coordinators was conducted to determine the presence and characteristics of sepsis performance improvement practices in acute care hospitals. The sampling frame was all non-federal acute care hospitals in the state (n=118 hospitals). The questionnaire (Supplement Appendix 1) was developed and refined by the study team, including an ED physician-administrator (AA), a quality engineer with a doctoral degree in epidemiology (JM), a critical care physician and health services researcher (NM), and research staff conducting surveys (TB, MB, AK). The instrument consisted of both structured questions and semi-structured prompts, intended to collect diverse responses representing the breadth of health care delivery, without restricting our data to findings expected by the study team. The questionnaire was field tested in two institutions for face validity and flow. Two trained research team members conducted the survey (TB and AK), and standardized prompts and a survey script were used to ensure uniformity across respondents. The survey questionnaire was designed to assess for the presence and characteristics of performance improvement practices in eight categories: sepsis committee, sepsis coordinator, physician sepsis champion, sepsis case review process, code sepsis response team, standardized process for sepsis patient identification, sepsis training, and sepsis registry. Survey respondents were identified by contacting hospitals via telephone and asking for the Director of Quality Improvement, if the hospital did not have this position, someone with a similar role was identified via speaking with hospital staff. If a respondent did not know the answer to a particular survey question, follow-up calls were scheduled with other members of the hospital team. Definitions used for the survey process are included in the Supplement Appendix 2. Data were collected using Research Electronic Data Capture software (REDCap) (Vanderbilt University, Nashville, TN).

### Data Sources and Definitions

Survey data were linked to two data sources: CMS SEP-1 score data and statewide hospital and hospital administrative claims data. SEP-1 score data were obtained from the CMS Hospital Compare public website for the reporting periods of April 2018 to March 2019.(16) State hospital association data containing administrative claims for all adults (18 years) treated in a hospital within the state between 2015 to 2019 were used to estimate sepsis risk-adjusted mortality by hospital.(17) Both datasets were deterministically linked using a unique hospital identifier to the telephone survey data. Hospital characteristics additionally included annual sepsis volume (from claims data), critical access hospital status (from CMS data), and rurality of hospital (from the zip-code tabulation area approximation of Rural-Urban Commuting Areas [RUCA] codes).(18)

### Key Measures & Outcomes

The primary exposure in this analysis was sepsis performance improvement activities, and the primary outcome in this study was hospital SEP-1 reporting status. Hospital SEP-1 reporting status was defined by presence of any SEP-1 scores during the reporting periods. Performance improvement measures were obtained from the study survey (Appendix 1). These measures were evaluated as binary (present/absent) for the primary analysis with characteristics of the components assessed in secondary analyses.

Secondary outcomes included SEP-1 adherence and hospital-specific risk-adjusted mortality. SEP-1 scores are a percentage from 0 to 100 and represent the proportion of eligible sepsis cases with complete adherence with both (1) a 3-hour bundle of serum lactate measurement, fluid resuscitation, blood cultures, and broad-spectrum antibiotics and (2) a 6-hour bundle including repeat lactate measurement, vasopressor administration, and reassessment of volume status/tissue perfusion. SEP-1 scores were retained as both a continuous measure and categorized by quartile (e.g. top quartile is 75th-99th percentile) of hospitals.

Risk-adjusted sepsis mortality was defined as hospital-specific observed-to-expected (O:E) sepsis mortality, assigning transferred cases to the first hospital. All adult patients presenting to the ED or inpatient unit with sepsis, fed by the implicit definition combining at least one infection diagnosis and one organ failure/dysfunction diagnosis code, were included in the mortality calculation. (17, 19) Expected mortality was calculated by estimating predicted probabilities for mortality for each case using a multivariable logistic regression model (age, race, sex, year, Elixhauser comorbidities (20), patient rurality, infection source, sepsis ICD-10-CM codes, palliative care, and organ dysfunction) to generate a predicted probability of in - hospital mortality. The O:E ratio was then calculated as the ratio of the sum of observed mortality divided by the sum of the expected probability of mortality for each hospital.

### Data Analysis

Characteristics of hospitals reporting SEP-1 data were compared to hospitals not reporting SEP-1 data using descriptive statistics and chi-square tests. Chi-squared and Fisher’s exact tests were used to compare characteristics of sepsis performance improvement practices by SEP-1 reporting status. SEP-1 scores and O:E mortality ratios were compared by sepsis performance improvement practices with Wilcoxon Mann Whitney test and across quartiles (gamma). Logistic and linear regression were used to identify associations between sepsis performance improvement practices and top- and bottom-preforming hospitals (defined as top and bottom 25% of SEP-1 hospitals, respectively) and SEP-1 scores, respectively. Quartiles were used to screen for threshold effects for high-performing hospitals, and then the continuous outcome was used to assess for associations between performance improvement practices and SEP-1 scores. For linear regression, the assumptions of the exposure-outcome relationship and homoskedasticity of residuals were assessed visualizing the pattern of observed versus predicted residuals. For the secondary outcome of O:E mortality ratios, a log-transformed model was used to satisfy these assumptions. Models adjusting for rurality of the hospital and critical access status were considered, but when relationships between rurality and critical access status and the outcome were not observed, only unadjusted models were presented. Statistical tests were considered significant at the p<0.05 threshold for two-tailed tests; analyses were conducted in SAS (version 9.4; SAS Institute, Cary, NC).

### Post-Hoc Sensitivity and Stratified Analyses

A stratified analysis assessed for differences in the relationship between performance improvement practices and sepsis mortality by SEP-1 reporting status. Two subcomponents (case review results reported to provider and results used in sepsis care plans) were hypothesized to be independently related with outcomes, so regression models estimated the association of these specific practices with outcomes. Last, a sensitivity analysis was performed assigning hospital outcomes based on last hospital for transferred patients (rather than first hospital).(21)

## Results

### Description of Study Population

All hospitals responded to the telephone survey (response rate 100%, N=118). Overall, hospitals had a median sepsis volume of 121 sepsis patients per year, 22% of hospitals were urban and 70% were rural critical access hospitals (Table 1). Forty-four hospitals (37%) reported SEP-1 adherence and the median SEP-1 score was 56.5 (IQR: 44.5 to 71.5). SEP-1 reporting hospitals were more likely to be urban (48% vs. 7%; %diff: 41%, 95%CI: 24 to 56%) and less likely to be critical access hospitals (23% vs. 97%; %diff: -74%, 95%CI: -84 to -58%) when compared to hospitals that did not report SEP-1 metrics. Top-performing hospitals (i.e. those in the top quartile) had SEP-1 scores between 73 and 97.

### Comparison of Performance Improvement Practices by SEP-1 Reporting Status

Overall, the majority of hospitals had a formal sepsis case identification process (92%, n=109), sepsis case review (75%, n=89), and sepsis training (66%, n=77). Other sepsis performance improvement practices were less common, including having a sepsis committee (39%, n=46) and a code sepsis response team (13%, n=15). Seven of the eight sepsis performance improvement components: sepsis committee (73 vs. 19%; difference 54%, 95%CI: 36 to 67%), sepsis coordinator (61 vs. 37%; difference 24%, 95%CI: 5 to 40%), physician sepsis champion (66 vs. 31%; difference 35%, 95%CI: 16 to 50%), sepsis case review process (89 vs. 68%; difference 21%, 95%CI: 5 to 34%), code sepsis response team (23 vs. 7%,; difference 16%, 95%CI: 3 to 31%), sepsis training (89 vs. 51%; difference 38%, 95%CI: 21 to 51%), and sepsis registry (66 vs. 26%; difference 40%, 95%CI: 22 to 55%) were more commonly reported in SEP-1 reporting hospitals (Figure 1). There was no difference in the use of a standardized process for sepsis patient identification between SEP-1 reporting and non-reporting hospitals.

Among hospitals that reported each sepsis performance improvement practice, we assessed differences in the characteristics of how the performance improvement practice was performed (Table S1). Overall, most hospitals performed the improvement practices similarly. For example, the 46 hospitals with sepsis committees, most practices were similar except more SEP-1 hospitals reported decreasing sepsis mortality as a committee goal and tracked sepsis mortality and bundle adherence. Sepsis committees were commonly chaired by clinicians or nurses with members including physicians, hospital administrators, pharmacists, nurses, and quality/safety specialists; most met monthly or quarterly and tracked sepsis mortality and sepsis bundle adherence. There were also no differences in the components of the code sepsis response teams among hospitals with a team, with teams being staffed 24 hours daily with physicians and nurses, activated by SIRS criteria or clinician concern, and being utilized hospital-wide.

However, there were differences in practices by SEP-1 reporting status for sepsis coordinators, physician sepsis champions, sepsis case review, sepsis case identification, sepsis training, and sepsis registry (Figure 2A-B)(Table S1).

### Association of Performance Improvement Practices with SEP-1 Scores

There was no association observed between sepsis performance improvement practices and low-performance (i.e. bottom-quartile) SEP-1 hospitals (Figure 3) (Table S2). There remained no association when the SEP-1 outcome was redefined as a continuous outcome or high-performance (i.e. top-quartile) SEP-1 hospitals.

### Association of Performance Improvement Practices with Mortality

Presence of a sepsis registry was associated with decreased odds of being in the bottom quartile of sepsis mortality (OR: 0.37; 95%CI 0.14 – 0.96, p=0.041) (Figure 3) (Table S3). When mortality was analyzed as a continuous outcome or as high-performance hospitals, these effects were no longer observed (Table S3). In this continuous model, presence of a sepsis committee was associated with lower hospital-specific mortality (log-transformed O:E ratio: -0.11; 95%CI -0.20 to -0.01).

### Association of Subcomponents with SEP-1 scores and Mortality

Two subcomponents of the case review process were independently assessed for associations with sepsis outcomes. Case review results reported by providers was not associated with SEP-1 scores (−0.43, 95%CI -2.75 – 1.88, p=0.708) or mortality (−0.03, 95%CI -0.13 – 0.07, p=0.585). Case review results used for sepsis care plans was not associated with SEP-1 scores (−1.29, 95%CI -4.00 – 1.42, p=0.342), but was associated with improved mortality (−0.10, 95%CI -0.19 to -0.00, p = 0.049).

### Sensitivity Analyses

When we reallocated transferred sepsis cases to the last hospital where they were treated (rather than the first), the results were similar (Table S4).

## Discussion

In our survey of hospitals in a predominantly rural state, we found that hospitals that report SEP-1 adherence were more likely to use performance improvement practices than those hospitals that do not report SEP-1. Among those that reported SEP-1 adherence, we did not find an association between individual performance improvement practices and SEP-1 score, but we did find that use of a sepsis registry, having a multidisciplinary sepsis committee, and using results from individual sepsis case review to refine sepsis protocols were all associated with decreased probability of a hospital being in the bottom quartile of sepsis risk-adjusted mortality. Perhaps most interesting, we observed that sepsis performance improvement practices are heterogeneous across hospitals, with very different local performance improvement programs.

One apparent explanation for this relationship could be that increased regulatory pressure facing hospitals who report SEP-1 may drive commitment to sepsis quality improvement. Seven of the eight sepsis performance improvement components were more commonly reported in SEP-1 reporting hospitals, and these hospitals also used their sepsis performance data for more quality improvement activities.

A relevant factor in our study is the types of hospitals included in the state where the study was conducted. Many hospitals that do not report SEP-1 are critical access hospitals, federally qualified small rural hospitals that are structurally different from higher volume centers. This factor alone might influence the sepsis quality improvement because these hospitals have fewer resources to focus on sepsis quality, lower sepsis volume, and many sepsis patients are transferred to other hospitals for inpatient care.(20)

The relationship between performance improvement and mortality, however, suggests that there may be an important relationship between performance improvement and patient outcomes. The 3 factors associated with mortality all are related to the use of data for process improvement: having a sepsis registry, using data from the registry to improve protocols, and having a multidisciplinary sepsis committee. This observation is especially interesting because of the difficulty in collecting and maintaining accurate sepsis quality data—the SEP-1 measure is extremely complex and requires manual chart review. Diagnosis codes are imprecise, (30) timing of care is complicated (31), and the roster of clinicians responsible for providing sepsis care is large and heterogeneous.

This finding also highlights another important issue—sepsis quality measures exempt low-volume hospitals. While this is important from the perspective of measurement, it is also important if a lack of quality measurement leads to insufficient incentive for performance improvement. An important next step is to consider ways to include low-volume hospitals in sepsis performance measurement, as prior work has shown performance improvement yielded larger returns in urban vs. rural hospitals.(22) Previous studies have found that rural hospitals that report SEP-1 perform well, but any metric of rural sepsis quality is hampered by low rural hospital reporting.(23)

This study has several limitations. To address the impact of acute changes due to the COVID-19 pandemic, hospitals were asked to answer based on their practices prior to the pandemic (i.e. December 2019). All survey data were collected from May 2020 to July 2020, which meant any changes were recent enough to be apparent and recall bias should be minimal.

Another limitation was the temporal impact of changes over time on SEP-1 scores and mortality. However, the primary outcome (SEP-1 scores) occurred only 15 months before the survey, so we do not expect significant directional bias. Another limitation is the observational nature of our data collection, which limits our ability to draw causal conclusions. The ability to measure actual outcomes and heterogenous performance improvement practices over a series of health systems, though, mitigates that limitation. Finally, our sample was drawn from a single Midwestern state. Despite that, the generalizability of this study is broad: the state where the study was conducted reflects the national distribution of SEP-1 scores well; with a state average of 51% compared to the national average of 50% (29) and includes rural and urban hospitals with a range of sepsis volumes.

## Conclusion

In conclusion, hospitals reporting SEP-1 performance are engaged in more performance improvement activities that those that are not. Some of these activities, including an institutional sepsis registry, using sepsis quality data to change hospital protocols, and having a multidisciplinary sepsis are associated with decreased hospital risk-adjusted mortality, but a relationship with SEP-1 performance was not observed. This finding is important in understanding the role of quality measurement on the process of performance improvement.

Future research should be focused on broadening the reach and scope of performance improvement practices. Public and government organizations may encourage or incentivize these activities, and these interventions may be associated with improved outcomes. Most important, future work should seek to validate which of these performance improvement activities are most effective in improving outcomes, because hospitals would value an evidence-based framework for performance improvement.

## Data Availability

SEP-1 score data were obtained from the CMS Hospital Compare public website for the reporting periods of April 2018 to March 2019. State hospital association data containing administrative claims for all adults (18 years) treated in a hospital within the state between 2015 to 2019 were used to estimate sepsis risk-adjusted mortality by hospital.

