## Supplementary material for "Hospitals That Report Severe Sepsis and Septic Shock Bundle (SEP-1) Compliance Have More Structured Sepsis Performance Improvement": Tables and Figures

**List of Tables and Figures**

**Table 1.** Hospital characteristics by SEP-1 reporting status.

**Figure 1.** Sepsis performance improvement practices by SEP-1 reporting status.
**Figure 2.** Sepsis performance improvement sub-components that differ by SEP-1 reporting status.
**Figure 3.** Association of sepsis performance improvement with SEP-1 scores and O:E mortality ratios (predicting bottom quartile performance).

**Table 1.** Hospital characteristics by SEP-1 reporting status.

| | Overall (N=118) | | SEP-1 Reporting Hospitals (n=44) | | Non-SEP-1 Reporting Hospitals (n=74) | | |
|---|---|---|---|---|---|---|---|
| **Hospital Characteristic** | **N or Median** | **% or IQR** | **n or Median** | **% or IQR** | **n or Median** | **% or IQR** | **p-value** |
| **Sepsis Volume** (Sepsis Visits per Year) | 121 | 68 – 326 | 933 | 244 – 1,494 | 78 | 56 - 697 | <0.001 |
| **Rurality (RUCA)** | | | | | | | <0.001 |
| Urban | 26 | 22.0 | 21 | 47.7 | 5 | 6.8 | |
| Large Rural City/Town | 15 | 12.7 | 13 | 29.6 | 2 | 2.7 | |
| Small Rural City/Town | 50 | 42.4 | 8 | 18.2 | 42 | 56.8 | |
| Isolated Small Rural Town | 27 | 22.9 | 2 | 4.6 | 25 | 33.8 | |
| **Critical Access Hospital Status** | 82 | 69.5 | 10 | 22.7 | 72 | 97.3 | <0.001 |
| **SEP-1 Score** (Median, IQR) | - | - | 56.5 | 44.5 – 71.5 | - | - | - |

IQR=Interquartile range; RUCA= Rural-Urban Commuting Area

**Figure 1.** Sepsis performance improvement practices by SEP-1 reporting status.

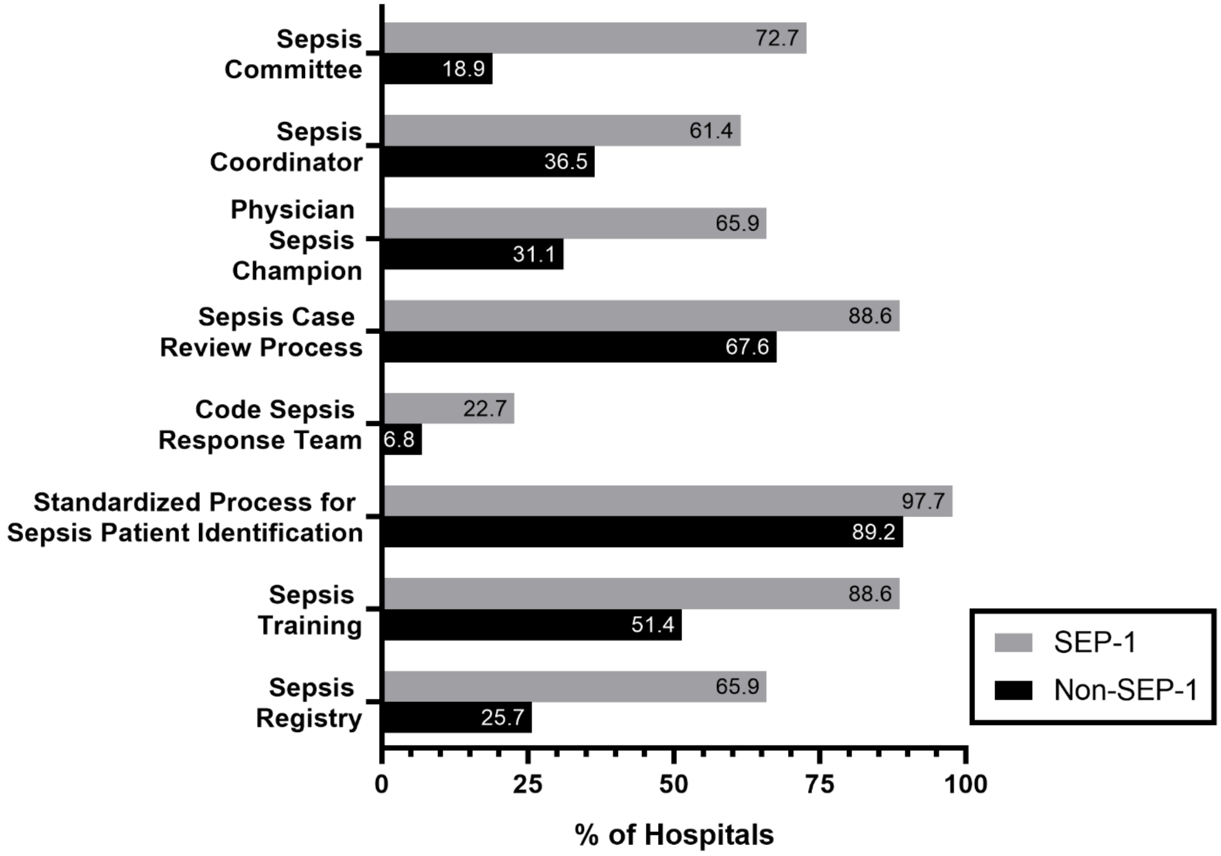

**Figure 2.** Sepsis performance improvement sub-components that differ by SEP-1 reporting status.

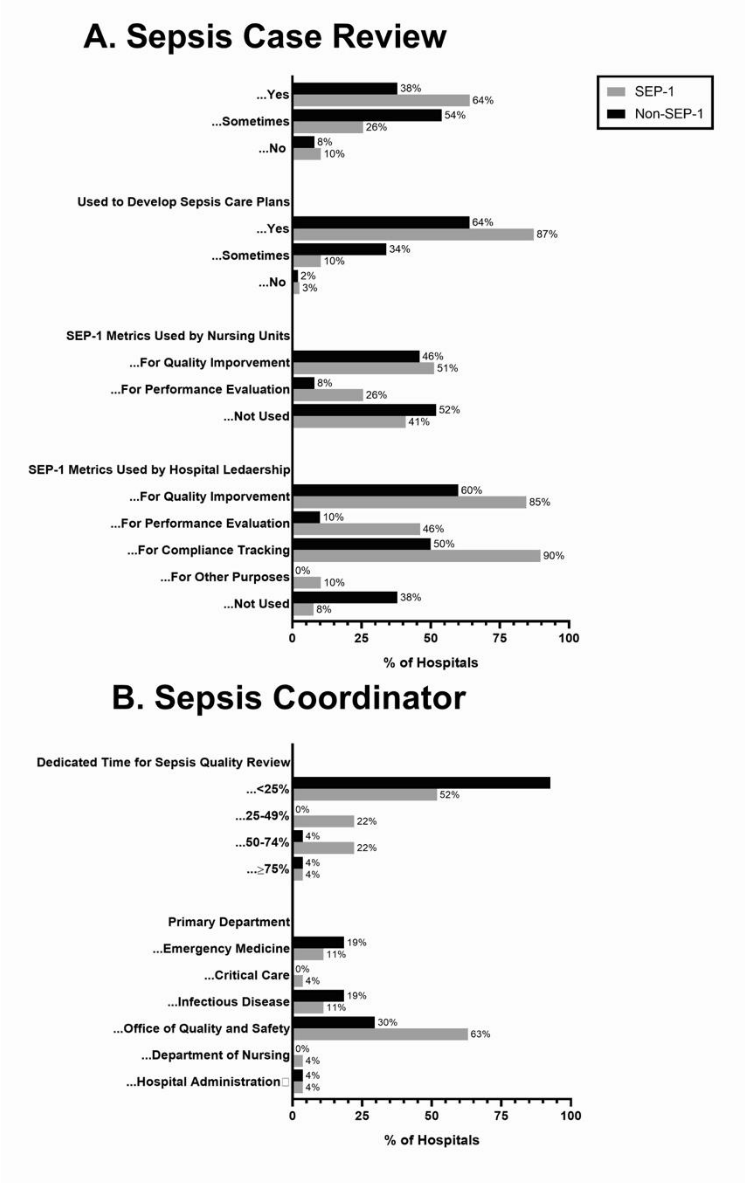

**Figure 3.** Association of sepsis performance improvement with SEP-1 scores and O:E mortality ratios (predicting bottom quartile performance).

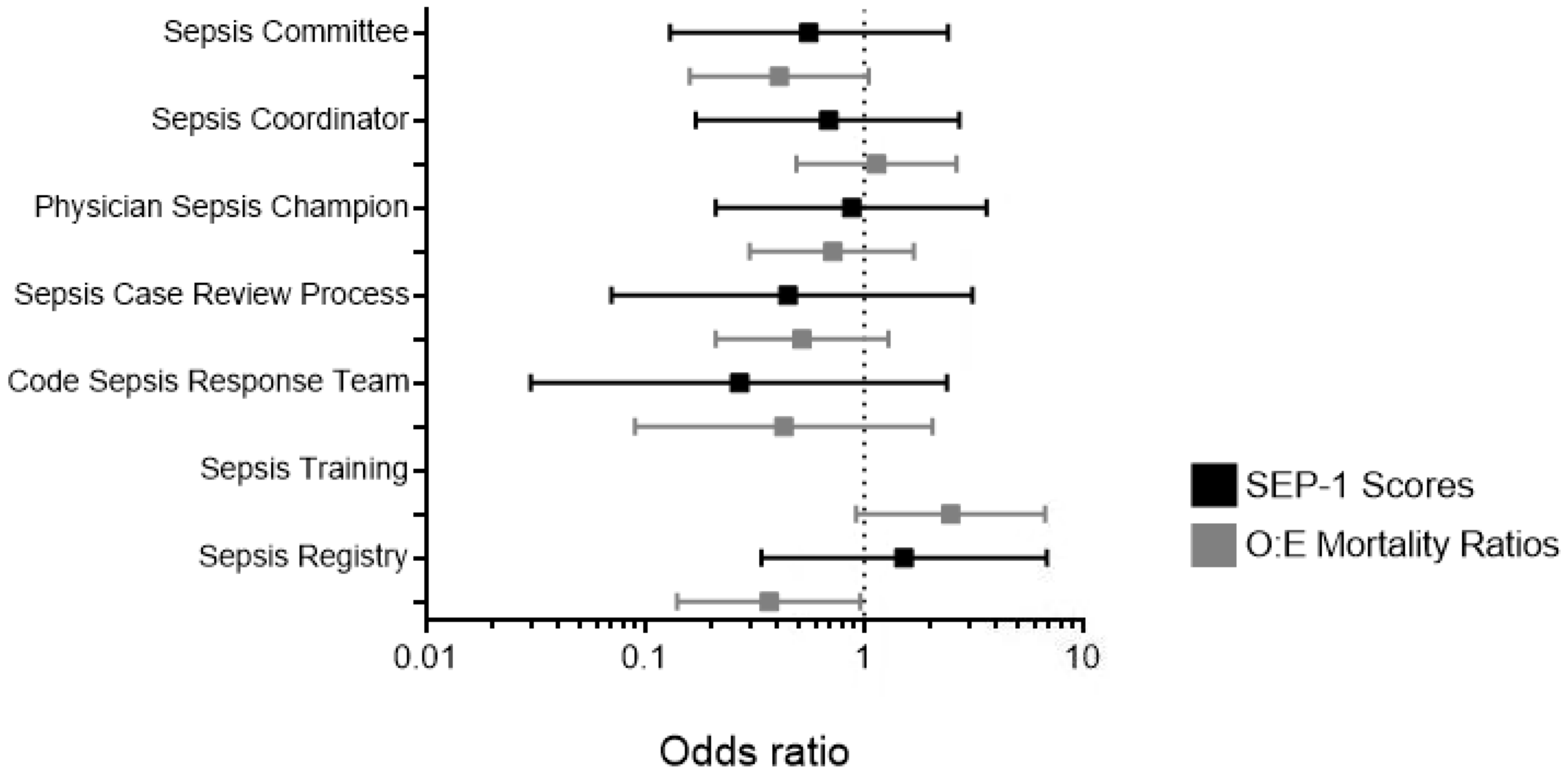


Since all top-performing hospitals (n=11) and most SEP-1 reporting hospitals had standardized processes for sepsis identification and sepsis training, the odds of top-quartile performance for these performance improvement practices could not be estimated.

**Supplementary Material List of Tables and Figures**

**Table S1.** Sepsis performance improvement practices by SEP-1 reporting status.
**Table S2.** Association of sepsis performance improvement practices with SEP-1 adherence.
**Table S3.** Association of sepsis performance improvement practices with hospital sepsis mortality.
**Table S4.** Sensitivity Analysis: Association of sepsis performance improvement practices with hospital sepsis mortality (O:E ratio is calculated to include sepsis patients transferred in to the hospital).

**Appendix 1.** Study survey questionnaire and script.
**Appendix 2.** Survey definitions.

**Table S1.** Sepsis performance improvement practices by SEP-1 reporting status.

| | Overall (N=118) | | SEP-1 Reporting Hospitals (n=44) | | Non-SEP-1 Reporting Hospitals (n=74) | | p-value |
|---|---|---|---|---|---|---|---|
| **SEPSIS COMMITTEE** | N | % | n | % | n | % | |
| **Have a Sepsis Committee (Yes/No)** | **46** | **39.0** | **32** | **72.7** | **14** | **18.9** | **<0.001** |
| Role of Sepsis Committee | | | | | | | |
| Reviews cases | 44 | 95.7 | 31 | 96.9 | 13 | 92.9 | 0.521 |
| Provide care recommendations | 40 | 87.0 | 29 | 90.6 | 11 | 78.6 | 0.350 |
| Improve performance improvement measures (e.g. SEP-1 scores) | 40 | 87.0 | 30 | 93.4 | 10 | 71.4 | 0.060 |
| Decrease Sepsis Mortality | 39 | 84.8 | 30 | 93.4 | 9 | 64.3 | 0.020 |
| Other | 4 | 8.7 | 2 | 6.3 | 2 | 14.3 | 0.575 |
| Committee Chair | | | | | | | 0.422 |
| Clinician | 14 | 30.4 | 10 | 31.3 | 4 | 28.9 | |
| Nurse | 13 | 28.3 | 8 | 25.0 | 5 | 35.7 | |
| Hospital Administrator | 6 | 13.0 | 6 | 18.8 | 0 | 0.0 | |
| Quality/Safety Specialist | 4 | 8.7 | 3 | 9.4 | 1 | 7.1 | |
| No Chair | 1 | 2.2 | 1 | 3.1 | 0 | 0.0 | |
| Other (Incl. multiple roles) | 8 | 17.4 | 4 | 12.5 | 4 | 12.5 | |
| Committee Members (Yes/No) | | | | | | | |
| Physician (MD/DO) | 43 | 93.5 | 31 | 96.9 | 12 | 85.7 | 0.216 |
| Physician Assistant | 3 | 6.5 | 2 | 6.3 | 1 | 7.1 | 0.999 |
| Pharmacist | 33 | 71.7 | 22 | 68.8 | 11 | 78.6 | 0.724 |
| Nurse Practitioner | 10 | 21.7 | 8 | 25.0 | 2 | 14.3 | 0.699 |
| Nurse | 43 | 93.5 | 29 | 90.6 | 14 | 100.0 | 0.543 |
| Hospital Administrator | 38 | 82.6 | 27 | 84.4 | 11 | 78.6 | 0.684 |
| Quality/Safety Specialist | 43 | 93.5 | 31 | 96.7 | 12 | 85.7 | 0.216 |
| Departments/Specialties Represented | | | | | | | |
| Emergency Medicine | 42 | 91.3 | 29 | 90.6 | 13 | 92.9 | 0.999 |
| Critical Care | 24 | 52.2 | 18 | 56.3 | 6 | 42.9 | 0.525 |
| Infectious Disease | 26 | 56.5 | 15 | 46.7 | 11 | 78.6 | 0.046 |
| Hospital Epidemiology | 22 | 47.8 | 16 | 50.0 | 6 | 42.9 | 0.754 |
| Pediatrics | 3 | 6.5 | 2 | 6.3 | 1 | 7.1 | 0.999 |
| Lab | 14 | 30.4 | 9 | 28.1 | 5 | 35.7 | 0.731 |
| Information Technology/Informatics | 8 | 17.4 | 6 | 18.8 | 2 | 14.3 | 0.999 |
| Metrics Tracked | | | | | | | |
| Sepsis Mortality | 39 | 84.8 | 30 | 93.8 | 9 | 64.3 | 0.020 |
| SEP-1 Scores or Other Sepsis Bundle Adherence | 40 | 87.0 | 31 | 96.9 | 9 | 64.3 | 0.007 |
| Components of Sepsis Bundle (Components only) | 6 | 13.0 | 3 | 9.4 | 3 | 21.4 | 0.350 |
| Other | 23 | 50.0 | 14 | 43.8 | 9 | 64.3 | 0.200 |

| | | | | | | | |
|---|---|---|---|---|---|---|---|
| Meeting Frequency | | | | | | | 0.141 |
| Weekly | 1 | 2.2 | 0 | 0.0 | 1 | 7.1 | |
| Twice per Month | 3 | 6.5 | 2 | 6.3 | 1 | 7.1 | |
| Monthly | 19 | 41.3 | 16 | 50.0 | 3 | 21.4 | |
| Quarterly | 22 | 47.8 | 13 | 40.6 | 9 | 64.3 | |
| As Needed | 1 | 2.2 | 1 | 3.1 | 0 | 0.0 | |
| **SEPSIS COORDINATOR** | | | | | | | |
| **Have a Sepsis Coordinator (Yes/No)** | **54** | **45.8** | **27** | **61.4** | **27** | **36.5** | **0.009** |
| Job Responsibilities | | | | | | | |
| Quality Improvement | 54 | 100.0 | 27 | 100.0 | 27 | 100.0 | 0.999 |
| Training/Education Requirement Checks | 29 | 53.7 | 18 | 66.7 | 11 | 40.7 | 0.056 |
| Conducting Training & Education | 36 | 66.7 | 21 | 77.8 | 15 | 55.6 | 0.083 |
| Guideline Implementation & Review | 45 | 83.3 | 21 | 77.8 | 24 | 88.9 | 0.467 |
| Abstracting Records for performance improvement Reporting | 43 | 79.6 | 20 | 74.1 | 23 | 85.2 | 0.311 |
| Bedside Practice Recommendations | 2 | 3.7 | 0 | 0.0 | 2 | 7.4 | 0.491 |
| Coordinator’s Other Roles | | | | | | | |
| Clinician | 19 | 35.2 | 9 | 33.3 | 10 | 37.0 | 0.776 |
| Hospital Administrator | 16 | 29.6 | 8 | 29.6 | 8 | 29.6 | 0.999 |
| Quality/Safety Specialist | 47 | 87.0 | 24 | 88.9 | 23 | 85.2 | 0.999 |
| Dedicated Time for Sepsis Quality Review | | | | | | | 0.001 |
| <25% | 39 | 72.2 | 14 | 51.9 | 25 | 92.6 | |
| 25-49% | 6 | 11.1 | 6 | 22.2 | 0 | 0.0 | |
| 50-74% | 7 | 13.0 | 6 | 22.2 | 1 | 3.7 | |
| ≥75% | 2 | 3.7 | 1 | 3.7 | 1 | 3.7 | |
| Dedicated Time for Direct Staff Education | | | | | | | 0.236 |
| <25% | 0 | 0.0 | 0 | 0.0 | 0 | 0.0 | |
| 25-49% | 0 | 0.0 | 0 | 0.0 | 0 | 0.0 | |
| 50-74% | 1 | 1.9 | 1 | 3.7 | 0 | 0.0 | |
| ≥75% | 50 | 92.6 | 26 | 96.3 | 24 | 88.9 | |
| Involved in other sepsis committees or activities (Yes/No) | 14 | 25.9 | 7 | 25.9 | 7 | 25.9 | 0.999 |
| Primary Department* | | | | | | | |
| Emergency Medicine | 8 | 14.8 | 3 | 11.1 | 5 | 18.5 | 0.704 |
| Critical Care | 1 | 1.9 | 1 | 3.7 | 0 | 0.0 | 0.999 |
| Infectious Disease | 8 | 14.8 | 3 | 11.1 | 5 | 18.5 | 0.704 |
| Office of Quality and Safety | 25 | 46.3 | 17 | 63.0 | 8 | 29.6 | 0.014 |
| Department of Nursing | 1 | 1.9 | 1 | 3.7 | 0 | 0.0 | 0.999 |
| Hospital Administration | 2 | 3.7 | 1 | 3.7 | 1 | 3.7 | 0.999 |
| Coordinator Training/Background | | | | | | | |

| | | | | | | | |
|---|---|---|---|---|---|---|---|
| Physician (MD/DO) | 0 | 0.0 | 0 | 0.0 | 0 | 0.0 | - |
| Physician Assistant | 1 | 1.9 | 0 | 0.0 | 1 | 3.7 | 0.999 |
| Pharmacist | 0 | 0.0 | 0 | 0.0 | 0 | 0.0 | - |
| Nurse Practitioner | 2 | 3.7 | 1 | 3.7 | 1 | 3.7 | 0.999 |
| Nurse | 51 | 94.4 | 26 | 96.3 | 25 | 92.6 | 0.999 |
| Hospital Administrator | 3 | 5.6 | 3 | 11.1 | 0 | 0.0 | 0.236 |
| Quality/Safety Specialist | 6 | 11.1 | 5 | 18.5 | 1 | 3.7 | 0.192 |
| Epidemiologist | 0 | 0.0 | 0 | 0.0 | 0 | 0.0 | - |
| Other | 1 | 1.9 | 0 | 0.0 | 1 | 3.7 | 0.999 |
| **PHYSICIAN SEPSIS CHAMPION** | | | | | | | |
| **Have a Physician Sepsis Champion (Yes/No)** | **52** | **44.1** | **29** | **65.9** | **23** | **31.1** | **<0.001** |
| Department | | | | | | | |
| Emergency Medicine | 23 | 44.2 | 12 | 41.4 | 11 | 47.8 | 0.642 |
| Critical Care | 4 | 7.7 | 3 | 10.3 | 1 | 4.4 | 0.621 |
| Infectious Disease | 6 | 11.5 | 3 | 10.3 | 3 | 13.0 | 0.999 |
| Hospital Epidemiology | 11 | 21.2 | 8 | 27.6 | 3 | 13.0 | 0.308 |
| Administration | 6 | 11.5 | 4 | 13.8 | 2 | 8.7 | 0.682 |
| Other | 6 | 11.5 | 1 | 3.5 | 5 | 21.7 | 0.076 |
| Other Formal Roles | | | | | | | |
| No Other Roles (FT Sepsis Quality) | 0 | 0.0 | 0 | 0.0 | 0 | 0.0 | - |
| Clinician | 44 | 84.6 | 25 | 86.2 | 19 | 82.6 | 0.999 |
| Hospital Administrator | 31 | 59.6 | 19 | 65.5 | 12 | 52.2 | 0.330 |
| Quality/Safety Specialist | 3 | 5.8 | 1 | 3.5 | 2 | 8.7 | 0.578 |
| Infection Control | 4 | 7.7 | 2 | 6.9 | 2 | 8.7 | 0.999 |
| Type of Work | | | | | | | |
| Clinical | 25 | 48.1 | 16 | 55.2 | 9 | 39.1 | 0.278 |
| Administration | 21 | 40.4 | 14 | 48.3 | 7 | 30.4 | 0.259 |
| Training | 37 | 71.2 | 23 | 79.3 | 14 | 60.9 | 0.218 |
| Quality | 34 | 65.4 | 19 | 65.5 | 15 | 65.2 | 0.982 |
| Other | 3 | 5.8 | 2 | 6.9 | 1 | 4.4 | 0.999 |
| Compensated for Sepsis Champion Role (Yes/No) | 8 | 15.4 | 8 | 27.6 | 0 | 0.0 | 0.006 |
| **SEPSIS CASE REVIEW** | | | | | | | |
| **Have a Sepsis Case Review Process (Yes/No)** | **89** | **75.4** | **39** | **88.6** | **50** | **67.6** | **0.010** |
| Frequency of Review | | | | | | | 0.061 |
| As Needed | 4 | 4.5 | 1 | 2.6 | 3 | 6.0 | |
| Daily | 7 | 7.9 | 3 | 7.7 | 4 | 8.0 | |
| Weekly | 14 | 15.7 | 8 | 20.5 | 6 | 12.0 | |
| Monthly | 49 | 55.1 | 16 | 41.0 | 33 | 66.0 | |
| Quarterly | 14 | 15.7 | 10 | 25.6 | 4 | 8.0 | |
| Other | 1 | 1.1 | 1 | 2.6 | 0 | 0.0 | |

| | | | | | | | |
|---|---|---|---|---|---|---|---|
| Sepsis Case Definition for Review | | | | | | | |
| Sepsis-Assoc.** Hospital Admissions | 60 | 67.4 | 23 | 59.0 | 37 | 74.0 | 0.134 |
| Sepsis-Assoc.** ICU Admissions | 1 | 1.1 | 1 | 2.6 | 0 | 0.0 | 0.438 |
| Sepsis Identified in ED | 3 | 3.4 | 1 | 2.6 | 2 | 4.0 | 0.999 |
| Sepsis Alert/Code Sepsis Activations | 3 | 3.4 | 2 | 5.1 | 1 | 2.0 | 0.579 |
| Sepsis Deaths | 3 | 3.4 | 3 | 7.7 | 0 | 0.0 | 0.081 |
| SEP-1 Bundle Not Completed | 17 | 19.1 | 11 | 28.2 | 6 | 12.0 | 0.054 |
| Sepsis Transfers Into Hospital | 0 | 0.0 | 0 | 0.0 | 0 | 0.0 | - |
| Sepsis Readmissions | 2 | 2.3 | 1 | 2.6 | 1 | 2.0 | 0.999 |
| Other | 18 | 20.2 | 10 | 25.6 | 8 | 16.0 | 0.261 |
| Reviewers of Cases | | | | | | | |
| Hospital Administration | 75 | 84.3 | 32 | 82.1 | 43 | 86.0 | 0.612 |
| Clinicians | 66 | 74.2 | 30 | 76.9 | 36 | 72.0 | 0.599 |
| Quality/Safety Specialists | 82 | 92.1 | 35 | 89.7 | 47 | 94.0 | 0.459 |
| Other | 4 | 4.5 | 1 | 2.6 | 3 | 6.0 | 0.628 |
| Results Reported to Providers | | | | | | | 0.026 |
| Yes | 44 | 49.4 | 25 | 64.1 | 19 | 38.0 | |
| Sometimes | 37 | 41.6 | 10 | 25.6 | 27 | 54.0 | |
| No | 8 | 9.0 | 4 | 10.3 | 4 | 8.0 | |
| Used to Develop Sepsis Care Plans | | | | | | | 0.013 |
| Yes | 66 | 74.2 | 34 | 87.2 | 32 | 64.0 | |
| Sometimes | 21 | 23.6 | 4 | 10.3 | 17 | 34.0 | |
| No | 2 | 2.3 | 1 | 2.6 | 1 | 2.0 | |
| Used to Re-evaluate Performance | | | | | | | 0.484 |
| Yes | 72 | 80.9 | 32 | 82.1 | 40 | 80.0 | |
| Sometimes | 8 | 9.0 | 2 | 5.1 | 6 | 12.0 | |
| No | 9 | 10.1 | 5 | 12.8 | 4 | 8.0 | |
| Results Reported to Departments | | | | | | | 0.541 |
| Yes | 42 | 47.2 | 20 | 51.3 | 22 | 44.0 | |
| Sometimes | 3 | 3.4 | 2 | 5.1 | 1 | 2.0 | |
| No | 44 | 49.4 | 17 | 43.6 | 27 | 54.0 | |
| SEP-1/Sepsis Metrics Used by Nursing Units | | | | | | | |
| For QI | 43 | 48.3 | 20 | 51.3 | 23 | 46.0 | 0.621 |
| For Performance Evaluation | 14 | 15.7 | 10 | 25.6 | 4 | 8.0 | 0.023 |
| Not Used | 42 | 47.2 | 16 | 41.0 | 26 | 52.0 | 0.304 |
| SEP-1/Sepsis Metrics Used by Hospital Leadership | | | | | | | |
| For QI | 63 | 70.8 | 33 | 84.6 | 30 | 60.0 | 0.011 |
| For Performance Evaluation | 23 | 25.8 | 18 | 46.2 | 5 | 10.0 | <0.001 |
| For Compliance Tracking | 60 | 67.4 | 35 | 89.7 | 25 | 50.0 | <0.001 |
| For Other Purpose | 4 | 4.5 | 4 | 10.3 | 0 | 0.0 | 0.034 |

| | | | | | | | |
|---|---|---|---|---|---|---|---|
| Not Used | 22 | 24.7 | 3 | 7.7 | 19 | 38.0 | 0.001 |
| **CODE SEPSIS** | | | | | | | |
| **Have a Code Sepsis Response Team (Yes/No)** | **15** | **12.7** | **10** | **22.7** | **5** | **6.8** | **0.012** |
| Activation Criteria | | | | | | | |
| Vital Signs | 1 | 6.7 | 0 | 0.0 | 1 | 6.7 | 0.333 |
| Clinician Concern | 4 | 26.7 | 4 | 40.0 | 0 | 0.0 | 0.231 |
| Specific Diagnoses | 1 | 6.7 | 0 | 0.0 | 1 | 6.7 | 0.333 |
| SIRS Criteria | 10 | 66.7 | 6 | 60.0 | 4 | 80.0 | 0.600 |
| Other | 2 | 13.3 | 2 | 20.0 | 0 | 0.0 | 0.524 |
| Responders | | | | | | | |
| Physician (MD/DO) | 13 | 86.7 | 9 | 90.0 | 4 | 80.0 | 0.999 |
| Physician Assistant | 0 | 0.0 | 0 | 0.0 | 0 | 0.0 | - |
| Pharmacist | 10 | 66.7 | 7 | 70.0 | 3 | 60.0 | 0.999 |
| Nurse Practitioner | 1 | 6.7 | 0 | 0.0 | 1 | 20.0 | 0.333 |
| Nurse | 15 | 100.0 | 10 | 100.0 | 5 | 100.0 | - |
| Quality/Safety Specialist | 1 | 6.7 | 0 | 0.0 | 1 | 20.0 | 0.333 |
| Lab | 12 | 80.0 | 9 | 90.0 | 3 | 60.0 | 0.242 |
| Other | 6 | 40.0 | 4 | 40.0 | 2 | 40.0 | 0.999 |
| Same as Code Sepsis Response Team (Yes/No) | 8 | 53.3 | 4 | 40.0 | 4 | 80.0 | 0.282 |
| Units Using Code Sepsis Team | | | | | | | |
| Hospital-wide | 15 | 100.0 | 10 | 100.0 | 5 | 100.0 | - |
| ICUs | 0 | 0.0 | 0 | 0.0 | 0 | 0.0 | - |
| ED | 0 | 0.0 | 0 | 0.0 | 0 | 0.0 | - |
| Clinics | 0 | 0.0 | 0 | 0.0 | 0 | 0.0 | - |
| Code Sepsis Team Staffed 24/7 | | | | | | | |
| Yes | 15 | 100.0 | 10 | 100.0 | 5 | 100.0 | - |
| No | 0 | 0.0 | 0 | 0.0 | 0 | 0.0 | - |
| Sometimes | 0 | 0.0 | 0 | 0.0 | 0 | 0.0 | - |
| **SEPSIS CASE IDENTIFICATION** | | | | | | | |
| **Have a Standardized Process for Sepsis Patient Identification** | **109** | **92.4** | **43** | **97.7** | **66** | **89.2** | **0.091** |
| Have an EMR Flag for Possible Sepsis | 102 | 93.6 | 39 | 90.7 | 63 | 95.5 | 0.322 |
| Units Using EMR Sepsis Flag | | | | | | | |
| Hospital-wide | 80 | 78.4 | 30 | 76.9 | 50 | 79.4 | 0.771 |
| ICUs | 2 | 2.0 | 2 | 5.1 | 0 | 0.0 | 0.144 |
| ED | 22 | 21.6 | 9 | 23.1 | 13 | 20.6 | 0.771 |
| Clinics | 0 | 0.0 | 0 | 0.0 | 0 | 0.0 | - |
| Other | 4 | 3.9 | 3 | 7.7 | 1 | 1.6 | 0.155 |
| Trigger Responses | | | | | | | |
| Activates Response Team | 6 | 5.5 | 4 | 9.3 | 2 | 3.0 | 0.210 |
| Alerts Provider | 101 | 92.7 | 39 | 90.7 | 62 | 93.9 | 0.710 |

| | | | | | | | |
|---|---|---|---|---|---|---|---|
| Activates Nursing Protocol | 33 | 30.3 | 16 | 37.2 | 17 | 25.8 | 0.203 |
| Flagged for Case Review | 11 | 10.1 | 7 | 16.3 | 4 | 6.1 | 0.108 |
| BPA | 5 | 4.6 | 0 | 0.0 | 5 | 7.6 | 0.155 |
| Nothing | 1 | 0.9 | 0 | 0.0 | 1 | 1.5 | 0.999 |
| Other Response | 2 | 1.8 | 1 | 2.3 | 1 | 1.5 | 0.999 |
| Nurse-Initiated Therapy from Triggers | | | | | | | |
| I.V. | 43 | 39.5 | 19 | 44.2 | 24 | 36.4 | 0.414 |
| Monitor | 39 | 35.8 | 18 | 41.9 | 21 | 31.8 | 0.285 |
| Labs | 43 | 39.5 | 21 | 48.8 | 22 | 33.3 | 0.106 |
| Fluids | 36 | 33.0 | 17 | 39.5 | 19 | 28.8 | 0.244 |
| Lactate | 44 | 40.4 | 22 | 51.2 | 22 | 33.3 | 0.064 |
| Blood Cultures | 42 | 38.5 | 21 | 48.8 | 21 | 31.8 | 0.074 |
| Oxygen | 5 | 4.6 | 2 | 4.7 | 3 | 4.6 | 0.999 |
| None | 59 | 54.1 | 17 | 39.5 | 42 | 63.6 | 0.014 |
| Other Therapy | 6 | 5.5 | 1 | 2.3 | 5 | 7.6 | 0.400 |
| Non-EMR Screening Practices | | | | | | | |
| Paper Checklist | 24 | 22.0 | 10 | 23.3 | 14 | 21.2 | 0.801 |
| Nursing Screen | 9 | 8.3 | 4 | 9.3 | 5 | 7.6 | 0.737 |
| Other | 3 | 2.8 | 1 | 2.3 | 2 | 3.0 | 0.999 |
| None | 78 | 71.6 | 31 | 72.1 | 47 | 71.2 | 0.921 |
| Units Using Non-EMR Screen (n=31) | | | | | | | |
| Hospital-wide | 15 | 48.4 | 7 | 58.3 | 8 | 42.1 | 0.379 |
| ICUs | 1 | 3.2 | 1 | 8.3 | 0 | 0.0 | 0.387 |
| ED | 12 | 38.7 | 3 | 25.0 | 9 | 47.4 | 0.274 |
| Clinics | 0 | 0.0 | 0 | 0.0 | 0 | 0.0 | - |
| Other | 3 | 9.7 | 2 | 16.7 | 1 | 5.3 | 0.544 |
| Rapid Response Team Has a Formal Sepsis Screening Practice (Yes/No) | 61 | 56.0 | 30 | 69.8 | 31 | 47.0 | 0.019 |
| **SEPSIS TRAINING** | | | | | | | |
| **Hospital Offers Sepsis Training (Yes/No)** | **77** | **66.3** | **39** | **88.6** | **38** | **51.4** | **<0.001** |
| Training Type | | | | | | | |
| Online | 40 | 52.0 | 21 | 53.9 | 19 | 50.0 | 0.736 |
| In-Person | 53 | 68.8 | 29 | 74.4 | 24 | 63.2 | 0.289 |
| Annual | 41 | 53.3 | 22 | 56.4 | 19 | 50.0 | 0.573 |
| Sepsis-Specific | 58 | 75.3 | 31 | 79.5 | 27 | 71.1 | 0.391 |
| Uniform Across Departments | 42 | 54.6 | 25 | 64.1 | 17 | 44.7 | 0.088 |
| Other | 2 | 2.6 | 0 | 0.0 | 2 | 5.3 | 0.240 |
| Trainings are Required | | | | | | | |
| Physician (MD/DO) | 9 | 11.7 | 8 | 20.5 | 1 | 2.6 | 0.029 |
| Physician Assistant | 5 | 6.5 | 4 | 10.3 | 1 | 2.6 | 0.358 |
| Pharmacist | 2 | 2.6 | 2 | 5.1 | 0 | 0.0 | 0.494 |
| Nurse Practitioner | 8 | 10.4 | 6 | 15.4 | 2 | 5.3 | 0.263 |

| | | | | | | | |
|---|---|---|---|---|---|---|---|
| Nurse | 46 | 59.7 | 27 | 69.2 | 19 | 50.0 | 0.085 |
| Quality/Safety Specialist | 6 | 7.8 | 6 | 15.4 | 0 | 0.0 | 0.025 |
| All Employees | 9 | 11.7 | 3 | 7.7 | 6 | 15.8 | 0.310 |
| Not Required | 14 | 18.2 | 6 | 15.4 | 8 | 21.1 | 0.519 |
| Emergency Department Staff | 6 | 7.8 | 4 | 10.3 | 2 | 5.3 | 0.675 |
| Other | 11 | 14.3 | 9 | 23.1 | 2 | 5.3 | 0.026 |
| Training Required at Time of Employment | | | | | | | 0.014 |
| Required for Some | 24 | 31.2 | 15 | 38.5 | 9 | 23.7 | |
| Required for All | 24 | 31.2 | 16 | 41.0 | 8 | 21.1 | |
| Not Required | 27 | 35.1 | 8 | 20.5 | 19 | 50.0 | |
| **SEPSIS REGISTRY** | | | | | | | |
| **Hospital Maintains a List/Registry of Sepsis Patients for QI (Yes/No)** | **48** | **40.7** | **29** | **65.9** | **19** | **25.7** | **<0.001** |
| List/Registry Used For | | | | | | | |
| Performance Evaluation | 18 | 37.5 | 14 | 48.3 | 4 | 21.1 | 0.057 |
| QI | 47 | 97.9 | 28 | 96.6 | 19 | 100.0 | 0.413 |
| Compliance Tracking | 37 | 77.1 | 24 | 82.8 | 13 | 68.4 | 0.304 |
| SEP-1 Reporting | 25 | 52.1 | 23 | 79.3 | 2 | 10.5 | <0.001 |
| Education | 26 | 54.2 | 20 | 69.0 | 6 | 31.6 | 0.011 |
| Research | 1 | 2.1 | 1 | 3.5 | 0 | 0.0 | 0.999 |
| Not Used | 0 | 0.0 | 0 | 0.0 | 0 | 0.0 | - |
| Other Use | 4 | 8.3 | 3 | 10.3 | 1 | 5.3 | 0.533 |
| List Includes Patients NOT identified for SEP-1 Data Abstraction (Yes/No) | 38 | 79.2 | 21 | 72.4 | 17 | 89.6 | 0.276 |
| Eligibility for Sepsis List | | | | | | | |
| Sepsis Death | 1 | 2.1 | 0 | 0.0 | 1 | 5.3 | 0.396 |
| Failed SEP-1 or SEP-1 Component | 1 | 2.1 | 0 | 0.0 | 1 | 5.3 | 0.396 |
| Met Code Sepsis Criteria | 2 | 4.2 | 2 | 6.9 | 0 | 0.0 | 0.512 |
| All Sepsis Patients | 32 | 66.7 | 19 | 65.5 | 13 | 68.4 | 0.835 |
| Suspected Sepsis | 15 | 31.3 | 7 | 24.1 | 8 | 42.1 | 0.232 |
| Other | 3 | 6.3 | 3 | 10.3 | 0 | 0.0 | 0.267 |
| Entire Chart Abstracted for Quality Metrics (Yes/No) | 32 | 66.7 | 16 | 55.2 | 16 | 84.2 | 0.037 |
| Methods for Chart Abstraction | | | | | | | 0.999 |
| Electronic Capture (EMR) | 2 | 6.3 | 1 | 6.3 | 1 | 6.3 | |
| Manual Capture | 30 | 93.8 | 15 | 93.8 | 15 | 93.8 | |

*One respondent declared two primary departments, so total is greater than sample size; **Sepsis-associated includes suspected sepsis cases

**Table S2.** Association of sepsis performance improvement practices with SEP-1 adherence.

| Outcome: | Continuous SEP-1 Score (linear regression) | | | Bottom Quartile Performance (logistic regression) | | | Top Quartile Performance (logistic regression) | | |
|---|---|---|---|---|---|---|---|---|---|
| **SEPSIS performance improvement Practices** | B | 95%CI | p-value | OR | 95%CI | p-value | OR | 95%CI | p-value |
| Sepsis Committee | -0.54 | -3.11 to 2.03 | 0.673 | 0.56 | 0.13 to 2.42 | 0.437 | 0.56 | 0.13 to 2.42 | 0.437 |
| Sepsis Coordinator | -0.34 | -2.69 to 2.02 | 0.774 | 0.69 | 0.17 to 2.73 | 0.593 | 1.14 | 0.28 to 4.67 | 0.858 |
| Physician Sepsis Champion | -0.72 | -3.14 to 1.69 | 0.548 | 0.88 | 0.21 to 3.64 | 0.854 | 0.52 | 0.13 to 2.12 | 0.362 |
| Sepsis Case Review Process | 0.05 | -3.57 to 3.67 | 0.979 | 0.45 | 0.07 to 3.13 | 0.419 | 0.45 | 0.07 to 3.13 | 0.419 |
| Code Sepsis Response Team | 0.57 | -2.17 to 3.30 | 0.678 | 0.27 | 0.03 to 2.39 | 0.238 | 0.70 | 0.12 to 3.90 | 0.679 |
| Standardized Process for Sepsis Patient Identification | -1.72 | -9.40 to 5.97 | 0.655 | * | | | * | | |
| Sepsis Training | -0.58 | -4.20 to 3.03 | 0.746 | * | | | * | | |
| Sepsis Registry | 1.19 | -1.49 to 3.32 | 0.447 | 1.52 | 0.34 to 6.86 | 0.583 | 1.52 | 0.34 to 6.86 | 0.583 |

Continuous outcome: beta represents association with a 5-point increase in SEP-1 score (%adherence). *Unable to estimate – all top and bottom quartile hospitals have sepsis training and a standardized process for sepsis identification.

**Table S3.** Association of sepsis performance improvement practices with hospital sepsis mortality.

| | Continuous O:E Mortality Ratio (linear regression – log transformed) | | | Bottom Quartile Performance (logistic regression) | | | Top Quartile Performance (logistic regression) | | |
|---|---|---|---|---|---|---|---|---|---|
| **SEPSIS performance improvement Practices** | B | 95%CI | p-value | OR | 95%CI | p-value | OR | 95%CI | p-value |
| Sepsis Committee | -0.11 | -0.20 to -0.01 | 0.036 | 0.41 | 0.16 – 1.05 | 0.064 | 1.90 | 0.85 – 4.26 | 0.121 |
| Sepsis Coordinator | -0.00 | -0.10 – 0.10 | 0.964 | 1.14 | 0.49 – 2.65 | 0.755 | 1.50 | 0.67 – 3.34 | 0.321 |
| Physician Sepsis Champion | -0.06 | -0.16 – 0.04 | 0.216 | 0.72 | 0.30 – 1.69 | 0.444 | 1.19 | 0.53 – 2.64 | 0.677 |
| Sepsis Case Review Process | -0.07 | -0.18 – 0.04 | 0.227 | 0.52 | 0.21 – 1.29 | 0.158 | 1.37 | 0.52 – 3.59 | 0.523 |
| Code Sepsis Response Team | -0.07 | -0.21 – 0.08 | 0.356 | 0.43 | 0.09 – 2.05 | 0.291 | 1.79 | 0.58 – 5.48 | 0.310 |
| Standardized Process for Sepsis Patient Identification | 0.12 | -0.06 – 0.30 | 0.199 | * | | | 1.45 | 0.29 – 7.38 | 0.651 |
| Sepsis Training | 0.03 | -0.07 – 0.14 | 0.515 | 2.49 | 0.92 – 6.71 | 0.073 | 1.71 | 0.71 – 4.12 | 0.233 |
| Sepsis Registry | -0.09 | -0.19 – 0.00 | 0.059 | 0.37 | 0.14 – 0.96 | 0.041 | 2.03 | 0.90 – 4.54 | 0.087 |

Mortality O:E ratios are log-transformed. * Unable to estimate – all bottom quartile hospitals have a standardized process for sepsis identification.

**Table S4.** Sensitivity Analysis: Association of sepsis performance improvement practices with hospital sepsis mortality (O:E ratio is calculated to include sepsis patients transferred in to the hospital).

| | Continuous O:E Mortality Ratio (linear regression – log transformed) | | | Bottom Quartile Performance (logistic regression) | | |
|---|---|---|---|---|---|---|
| **SEPSIS Performance Improvement Practices** | B | 95%CI | p-value | OR | 95%CI | p-value |
| Sepsis Committee | -0.08 | -0.18 – 0.02 | 0.130 | 0.41 | 0.16 – 1.05 | 0.064 |
| Sepsis Coordinator | 0.02 | -0.08 – 0.12 | 0.625 | 1.37 | 0.59 – 3.18 | 0.459 |
| Physician Sepsis Champion | -0.04 | -0.14 – 0.06 | 0.385 | 0.59 | 0.25 – 1.41 | 0.234 |
| Sepsis Case Review Process | -0.05 | -0.17 – 0.06 | 0.387 | 0.52 | 0.21 - 1.29 | 0.158 |
| Code Sepsis Response Team | -0.05 | -0.20 – 0.10 | 0.525 | 0.43 | 0.09 – 2.05 | 0.291 |
| Standardized Process for Sepsis Patient Identification | 0.10 | -0.09 – 0.29 | 0.287 | * | | |
| Sepsis Training | 0.05 | -0.05 – 0.16 | 0.298 | 3.26 | 1.14 – 9.34 | 0.028 |
| Sepsis Registry | -0.10 | -0.20 to -0.00 | 0.048 | 0.37 | 0.14 – 0.96 | 0.041 |

Mortality O:E ratios are log-transformed.

**Appendix 1.** Study survey questionnaire and script.

**Facility ID:** **Hospital Name:**

**Introduction:**

Good (afternoon/morning), my name is Ty Bolte and I am a medical student calling on behalf of Dr. Ahmed at the University of Iowa Department of Emergency Medicine. Thank you for your time.

I'm working with Dr. Ahmed on a project to survey hospitals in (Florida OR Iowa) on current sepsis quality improvement practices. We hope that this project will help inform regional and state sepsis practices and determine which are most effective in improving SEP-1 scores and sepsis mortality.

We're looking for someone who understands the sepsis QI practices at your hospital and would be able to answer some questions regarding training and sepsis care. All the data you share will be kept strictly confidential and will not be linked to your hospital in any published reports – all findings will be reported in aggregate. The survey will take around 10 minutes, and your participation is extremely important in the success of this project.

Can you please help us?

**Please note that these practices should be reflective of your policies and procedures prior to the COVID-19 pandemic.**

**Questions regarding quality improvement:**

For this survey I'm going to ask you some questions about sepsis quality improvement and care at your institution.

1. Does your hospital have a sepsis committee? Tell me about the committee.
    a. What is the role of the committee?
        i. Review cases ☐
        ii. Provide sepsis care recommendations☐
        iii. Improve QI measures like SEP-1 scores ☐
        iv. Decrease sepsis mortality ☐
        v. Other ☐
            1. ________________________________________

b. Who is the chairperson of the committee?
    i. Clinician ☐
    ii. Nurse ☐
    iii. Hospital Administrator ☐
    iv. Quality and Safety Specialist ☐
    v. Other
        1. ________________________________________
c. Tell me about the types of providers that are on the team? Are there any specific specialties represented?
    i. Physician (MD, DO) ☐
    ii. Physician Assistant ☐
    iii. Pharmacist ☐
    iv. Nurse Practitioner ☐
    v. Nurse ☐
    vi. Quality and Safety Specialist ☐
    vii. Hospital Administrator ☐
    viii. Other ☐
        1. ________________________________________
    ix. Emergency Medicine ☐
    x. Critical Care ☐
    xi. Infectious Diseases ☐
    xii. Hospital Epidemiology ☐
    xiii. Pediatrics ☐
d. What types of metrics does your committee track?
    i. Sepsis mortality ☐
    ii. SEP-1 ☐
    iii. Other ☐
        1. ________________________________________
e. How often does the committee meet?
    i. Weekly ☐
    ii. Twice a month ☐

iii. Monthly ☐
iv. Quarterly ☐
v. Twice a year ☐
vi. Yearly ☐

2. Does your hospital have a sepsis coordinator?
    i. Yes ☐
    ii. No ☐
    a. Describe this person's job function.
        i. Quality improvement ☐
        ii. Ensuring training and education requirements are met ☐
        iii. Conducting training/education ☐
        iv. Guideline implementation and review ☐
        v. Abstracting sepsis records for quality reporting ☐
    b. Does this individual have other roles in your hospital?
        i. Clinician ☐
        ii. Hospital Administrator ☐
        iii. Quality and Safety Specialist ☐
    c. How much of this person's time is allocated to sepsis quality review? (quality improvement project, data review, etc.)
        i. Less than a quarter ☐
        ii. A quarter to half ☐
        iii. Half to three-fourths ☐
        iv. Three-fourths to full-time ☐
    d. What proportion of this person's time is spent providing direct staff education?
        i. Less than a quarter ☐
        ii. A quarter to half ☐
        iii. Half to three-fourths ☐
        iv. Three-fourths to full-time ☐
    e. Does this person coordinate or involved in other sepsis activities or committees outside of their position?
        i. Yes ☐

    ii. No ☐

f. Which department does this coordinator primarily work for?

    i. Emergency Medicine ☐
    ii. Critical Care ☐
    iii. Infectious Diseases ☐
    iv. Hospital Epidemiology ☐
    v. Office of Chief Medical Officer ☐
    vi. Office of Quality and Safety ☐
    vii. Department of Nursing ☐
    viii. Department of Pharmacy ☐

g. What types of training does this person have?

    i. Physician (MD, DO) ☐
    ii. Physician Assistant ☐
    iii. Pharmacist ☐
    iv. Nurse Practitioner ☐
    v. Nurse ☐
    vi. Quality and Safety Specialist ☐
    vii. Hospital Administrator ☐
    viii. Epidemiologist ☐
    ix. Other ☐
        1. ________________________________________

3. Does your institution have a physician sepsis champion?

    i. Yes ☐
    ii. No ☐

a. In what department?

    i. Emergency Medicine ☐
    ii. Critical Care ☐
    iii. Infectious Diseases ☐
    iv. Hospital Epidemiology ☐

b. Does that person have a formal role other than sepsis champion?
    i. No- they do full time sepsis quality activity ☐
    ii. Clinician ☐
    iii. Hospital Administrator ☐
    iv. Quality and Safety Specialist ☐
    v. Infection Control ☐

c. What kind of work does he/she do as sepsis champion?
    i. Clinical ☐
    ii. Administration ☐
    iii. Training ☐
    iv. Quality ☐

d. Does your hospital pay him/her to do that work?
    i. Yes ☐
    ii. No ☐

4. Does your hospital have a sepsis case-review process? Tell me more about this process.
    a. When and how are these case reviews completed?
        i. Weekly ☐
        ii. Twice a month ☐
        iii. Monthly ☐
        iv. Quarterly ☐
        v. Twice a year ☐
        vi. Yearly ☐
    b. Which sepsis patients qualify for case review?
        i. From all sepsis-associated admissions to the hospital ☐
        ii. From all sepsis-associated admissions to the ICU ☐
        iii. From all sepsis patients identified in the emergency department ☐
        iv. From all patients for whom a "sepsis alert" or "code sepsis" is activated ☐
        v. From all patients who die of sepsis ☐
        vi. From patients for who the SEP-1 bundle is not completed ☐

   vii. From patients who are transferred to your hospital with sepsis ☐
   viii. From patients who are readmitted after a sepsis-related discharge ☐
   ix. Other ☐

c. Who sees the results of case reviews?
   i. Hospital Administration ☐
   ii. Clinicians ☐
   iii. Quality and Safety Specialists ☐
d. Do you monitor performance for specific components of the Sepsis bundle?
   i. Yes ☐
   ii. No ☐
e. Are results reported back to providers involved in that patient's care? (physicians, nurses, etc.)
   i. Yes ☐
   ii. No ☐
   iii. Sometimes ☐
f. Are these data used to develop a plan for improvement in sepsis care?
   i. Yes ☐
   ii. No ☐
   iii. Sometimes ☐
g. Are these data used to re-evaluate performance using outcome-based measures?
   i. Yes ☐
   ii. No ☐
   iii. Sometimes ☐

5. Tell me about how you use SEP-1 data in your organization?
   a. Is SEP-1 compliance reported back to individual departments?
      i. Yes ☐
      ii. No ☐
      iii. Sometimes ☐
   b. Is it reported back to providers involved in that patient's care?

    i. Yes ☐
    ii. No ☐
    iii. Sometimes ☐

c. How is SEP-1 data used by nursing units?
    i. Used for quality improvement ☐
    ii. Used for performance evaluation ☐
    iii. Not used ☐

d. Are SEP-1 results reported in hospital leadership meetings? How are those data used?
    i. Used for performance evaluation ☐
    ii. Used for quality improvement ☐
    iii. Used for compliance tracking ☐
    iv. Used for other purpose ☐
    v. Not reported ☐

**Hospital Characteristics**

6. Does your hospital have a Code Sepsis response team? Tell me about this team. (*If "No", go to Question 7)*
    a. What criteria activates this team?
        i. Vital signs ☐
        ii. Clinician concern ☐
        iii. Specific diagnosis ☐
        iv. SIRS criteria ☐
        v. Other ☐
            1. ________________________________________
    b. Who responds with the team?
        i. Physician (MD, DO) ☐
        ii. Physician Assistant ☐
        iii. Pharmacist ☐
        iv. Nurse Practitioner ☐
        v. Nurse ☐
        vi. Quality and Safety Specialist ☐

vii. Other ☐
1. ________________________________________
c. Is this the same team as your Rapid Response Team (RRT)?
i. Yes ☐
ii. No ☐
d. What units use the code sepsis team?
i. Hospital-wide ☐
ii. Intensive Care Units ☐
iii. Emergency Department ☐
iv. Clinics ☐
e. Is the team staffed 24/7, every day?
i. Yes ☐
ii. No ☐
iii. Sometimes ☐

**Process for identification of septic patients:**

7. Does your hospital have a standardized process for the identification of septic patients? Tell me about this process?
a. Does your electronic medical record flag cases of possible sepsis?
i. Yes ☐
ii. No ☐
b. If so, where is it used
i. Hospital-wide ☐
ii. Intensive Care Units ☐
iii. Emergency Department ☐
iv. Clinics ☐
c. What happens when the trigger identifies a patient?
i. Activates a response team ☐
ii. Alerts provider ☐
iii. Activates a nursing protocol (IV, monitor, labs, fluids, etc.) ☐
iv. Flagged for case review ☐

v. Nothing ☐

d. Do nurses have a protocol to initiate therapy based on certain sepsis triggers without a physician order? If so, for what treatments?
   i. IV ☐
   ii. Monitor ☐
   iii. Labs ☐
   iv. Fluids ☐
   v. Lactate ☐
   vi. Blood cultures ☐
   vii. No ☐
e. Are there non-EMR based screening practices?
   i. Paper checklist ☐
   ii. Nursing screen ☐
   iii. Other ☐
      1. ________________________________________
   iv. No ☐
f. Where are the non-EMR based screening practices used?
   i. Hospital-wide ☐
   ii. Intensive Care Units ☐
   iii. Emergency Department ☐
   iv. Clinics ☐
g. Does the Rapid Response Team have a formal sepsis screening practice?
   i. Yes ☐
   ii. No ☐

**General**

8. Does your hospital offer sepsis training? Tell me about these trainings.
   i. Online ☐
   ii. In-person ☐
   iii. Annual ☐

    iv. Sepsis specific ☐
    v. Uniform across departments ☐
    vi. No trainings. ☐

a. Are any of these trainings required for clinicians/nursing staff?
    i. Required for Physicians (MD, DO) ☐
    ii. Required for Physician Assistants ☐
    iii. Required for Pharmacists ☐
    iv. Required for Nurse Practitioners ☐
    v. Required for Nurses ☐
    vi. Required for Quality and Safety Specialists ☐
    vii. Required for all employees ☐
    viii. Not required ☐

b. Required at the time of employment?
    i. Required for some ☐
    ii. Required for all ☐
    iii. Not required ☐

9. Does your hospital maintain a list/registry of sepsis patients for QI?
    i. Yes ☐
    ii. No ☐

a. What do you use the list for?
    i. Used for performance evaluation ☐
    ii. Used for quality improvement ☐
    iii. Used for compliance tracking ☐
    iv. Used for SEP-1 reporting ☐
    v. Used for education ☐
    vi. Used for research ☐
    vii. Not used ☐

b. Does that list include patients NOT identified for SEP-1 data abstraction?
    i. Yes ☐

ii. No ☐

c. How does a patient get added to the sepsis registry?
   i. Patients with Sepsis mortality ☐
   ii. Patients who failed SEP-1 measure/Patients who failed a component of SEP-1 ☐
   iii. Patients who met criteria for a Code Sepsis ☐
   iv. All patients who were diagnosed with sepsis ☐

d. If a patient is on the list, is the entire chart abstracted for quality metrics?
   i. Yes ☐
   ii. No ☐

e. If yes, how are these data obtained?
   i. Electronically captured in the EMR (automatic process) ☐
   ii. Through manual chart review ☐

10. What do you think is the biggest barrier to sepsis care at your hospital?
   i. Free text question, write responses
      1. ________________________________________

11. What do you think is the best part about sepsis care sepsis at your hospital?
   i. Free text question, write responses
      1. ________________________________________

Thank you for your time. Many participants are interested in our findings, so we are also collecting e-mail addresses so that we can share results and reports with you at the end of our project. Would you like to provide your email so that I can provide you with a summary of our findings once our project is concluded?

**Appendix 2.** Survey definitions.

**Sepsis Committee**: a formalized group that meets routinely with the goal to discuss sepsis care. The committee can have many functions, including; completing sepsis case reviews, providing sepsis care recommendations, improving compliance on CMS measures, decreasing sepsis mortality, etc.
**Sepsis committee chairperson**: This person is generally the “leader” of the committee or someone in a leadership role who facilitates the sepsis committee meetings and may have additional responsibilities related to that role. The official title may not be “sepsis committee chairperson” but fills a similar role.
**Sepsis Coordinator**: A sepsis coordinator is someone who is responsible for various job functions related to sepsis, generally for the whole hospital. This person may lead or coordinate many sepsis initiatives and may have other job functions as well. The official title may not be “sepsis coordinator” but fills a similar role.
**Sepsis champion**: This person is a staff member who has taken an additional role involving sepsis care or QI, generally in addition to their primary role and most likely just for their department/team. Official title may not be “Sepsis champion” but fills a similar role.
**Sepsis case review process**: a formal process in which sepsis cases are reviewed. This may be separate than the Sepsis committee, or it may be a part of it. It also may include SEP-1 patients, or these patients may be in a separate review.
**Sepsis bundle**: the criteria for the care that patients with sepsis must receive to be compliant in the CMS SEP-1 measure. Some hospitals track may track specific elements that are commonly missed.
**Code Sepsis response team**: a specialized team that can be activated to care for a sepsis patient. May have specialized training or activation criteria.
**Code Sepsis:** a standardized process that is used to alert providers or activate a sepsis response team. Criteria used to activate the code is variable.
**EMR sepsis flag**: an electronic flag that is generated in response to specific criteria that will alert providers to a case of possible sepsis.
**Sepsis Registry**: a list of patients, generally who had or were suspected to have sepsis, that hospitals may keep for various reasons including QI, performance evaluation, quality reporting, etc. This list may include patients who do not qualify for SEP-1 abstraction.
